# Latent transdiagnostic dimensions linking sleep and psychopathology map onto distinct connectome-wide networks

**DOI:** 10.64898/2026.09.20.26363508

**Authors:** Yishan Kou, Xueer Jin, Xu Lei, Debo Dong, Yulin Wang

## Abstract

Sleep disturbance is common and clinically consequential across mental disorders. Yet its transdiagnostic role remains difficult to interpret because sleep is often reduced to a global score and psychiatric symptoms to diagnosis-specific outcomes. Here, we leveraged the YaleNeuroConnect cohort to test whether subjective sleep components align with separable dimensions of symptom-related features and connectome-wide organization. We applied canonical correlation analysis to seven Pittsburgh Sleep Quality Index components and 55 clinical, affective, cognitive and interpersonal measures in 260 diagnostically diverse subjects, and mapped the resulting dimensions onto resting-state functional connectivity using network-based statistics. We identified two sleep-psychopathology dimensions. The dominant and cross-validated dimension linked widespread subjective sleep disturbance, especially daytime dysfunction, to general symptom burden, including global severity, perceived stress and fatigue. This dimension was expressed in reduced coupling between the default mode network and frontoparietal and cerebellar systems, reduced cerebellar-subcortical coupling, and stronger coupling between the somatomotor network and higher-order systems. A second, more circumscribed dimension linked shorter sleep duration and prolonged sleep latency to lower self-assurance and positive affect, together with heightened affective and perceptual sensitivity. This dimension showed stronger coupling between cerebellar and dorsal attention systems, together with weaker coupling between the default mode network and attention or cerebellar systems. These findings suggest that sleep is not merely a comorbid symptom, but a multidimensional organizing feature of transdiagnostic psychopathology. Distinguishing broad sleep-related distress from sleep-initiation and sensitivity profiles may help refine brain-based stratification and identify sleep dimensions relevant to intervention.

## Introduction

Sleep disturbances are highly prevalent across psychiatric disorders and frequently co-occur with a wide range of symptom dimensions [1–3]. Although they have traditionally been viewed as secondary features to specific diagnoses [4], accumulating evidence supports a transdiagnostic perspective [5–7], indicating that their associations with psychopathology are heterogeneous and multidimensional. This perspective aligns with dimensional models of psychopathology [8,9] such as the internalizing and externalizing framework, which capture processes shared across diagnostic categories [10]. Importantly, different psychiatric disorders exhibit distinct patterns of sleep disturbances, and no two disorders appear to share an identical sleep phenotype [11,12]. This observation argues against a unitary account and instead suggests that sleep disturbances may reflect shared vulnerability processes rather than disorder-specific features.

Sleep itself is inherently a multidimensional construct rather than a simple contrast between “good” and “poor” [13]. The Pittsburgh Sleep Quality Index (PSQI) provides a well-established multidimensional assessment of subjective sleep quality. However, prior research has predominantly relied on its global score, potentially overlooking meaningful heterogeneity across distinct sleep dimensions [14]. This issue is particularly relevant because psychiatric disorders are characterized by distinct sleep profiles, and no single sleep parameter is uniquely altered in any specific disorder [12]. Specifically, individual sleep dimensions such as duration and efficiency do not necessarily vary in parallel and may differ in clinical relevance. For instance, daytime dysfunction is more closely linked to sleep continuity [12,15,16], while metabolic syndrome shows stronger associations with sleep regularity [17].

Sleep health frameworks further underscore that sleep cannot be reduced to a single index of poor sleep. The Ru-SATED model, for example, conceptualizes sleep health across multiple dimensions, including regularity, satisfaction, alertness, timing, efficiency and duration [18]. Although the present study was based on the PSQI rather than a dedicated Ru-SATED assessment, the PSQI provides a clinically established measure that captures several core subjective dimensions of sleep, including perceived sleep quality, sleep latency, sleep duration, sleep efficiency, sleep disturbances, use of sleep medication and daytime dysfunction. This makes it well suited for testing whether separable aspects of subjective sleep covary with distinct transdiagnostic symptom profiles. Consistent with this multidimensional view, our previous work showed that distinct sleep-health domains covary with distributed intrinsic functional connectivity patterns, supporting the value of treating sleep as a multidimensional phenotype in brain-connectivity research [13].

There is a growing consensus that psychopathology is multidimensional rather than unitary [9,19,20]. Constructs such as general distress, negative affect and deficits in positive affect [21] cut across traditional diagnostic boundaries, suggesting that categorical diagnoses may obscure shared variance across disorders [22]. Although comorbidity is commonly observed within categorical diagnoses, a latent structure perspective suggests that these co-occurring diagnoses may arise from shared underlying dimensions [23]. Previous studies have further shown that transdiagnostic dimensions of psychopathology are linked to large-scale brain network organization [24]. Taken together, these developments suggest that a transdiagnostic framework may provide a more suitable basis than categorical diagnosis for modeling the relationship between sleep and psychopathology.

Most previous studies have examined associations between single sleep indices and a narrow set of symptoms. For example, shorter sleep duration is associated with greater psychological distress [25], whereas poorer overall sleep quality is linked with stress-related symptoms [26]. However, such one-to-one approaches have been increasingly questioned because they produce fragmented findings and often show limited reproducibility [27]. As a result, the existing literature remains difficult to integrate and offers only limited insight into how distinct aspects of sleep relate to the broader structure of psychopathology in a transdiagnostic framework. Therefore, it remains unclear which specific sleep features reliably co-vary with particular psychopathological dimensions, and whether these complex relationships can be captured by a lower-dimensional latent structure. To address this issue, multivariate latent-variable approaches, such as canonical correlation analysis (CCA), have been increasingly applied to identify robust patterns of covariation in psychiatric and brain-behavior research [28].

Accumulating evidence suggests that sleep disturbances are associated with alterations in resting-state functional connectivity (rsFC) and large-scale brain networks [13,29], including the default mode and subcortical systems [30,31]. Nevertheless, most neuroimaging studies remain constrained by reductionist designs that focus on single sleep parameters, selected brain regions, or a limited number of networks. These approaches often rely on case-control comparisons, reducing sensitivity to distributed, multivariate effects expressed across the connectome. This limitation is particularly important given that mental disorders are increasingly conceptualized as disorders of large-scale network organization rather than isolated regional abnormalities [32–34], and that transdiagnostic dimensions can map onto distinct connectome-wide patterns of connectivity [35,36]. Moreover, while latent-variable approaches can capture shared variance at the clinical level, they do not directly specify the underlying neurobiological architecture through which these dimensions are expressed. As a result, it remains unclear whether distinct latent dimensions linking sleep and psychopathology correspond to different connectome-wide network configurations.

In the present study, we addressed these questions in the transdiagnostic YaleNeuroConnect sample [37] by applying CCA to identify latent dimensions linking multidimensional sleep characteristics with transdiagnostic symptom-related features. We then tested whether these dimensions were associated with distinct connectome-wide patterns of resting-state functional connectivity. We hypothesized that sleep-psychopathology associations would not be captured by a single common factor, but would instead reflect separable dimensions linking specific sleep features with distinct symptom profiles and large-scale brain network configurations.

## Results

### Two significant latent dimensions linked multidimensional sleep and symptom-related features

Out of seven LCs, CCA identified two significant dimensions of covariation between sleep and symptom-related measures after FDR correction [Dimension 1 canonical correlation = 0.84, p = 9.9 × 10^-5 (Fig. 1a); Dimension 2 canonical correlation = 0.62, p = 8.0 × 10^-4 (Fig. 2a)]. The two dimensions explained 56.6% and 14.2% of the covariance between the sleep and symptom-related data, respectively. LC1 represented a broad sleep-symptom pattern, capturing coordinated variation across multiple sleep and symptom measures. LC2 reflected a more specific covariance pattern centered on shorter sleep duration, prolonged sleep latency, and sensitivity-related symptom features. Cross-validation supported LC1 but not LC2, indicating that LC1 captured a more robust and generalizable sleep-symptom dimension, whereas LC2 represented a more circumscribed dimension that should be interpreted cautiously.

**Fig. 1.**
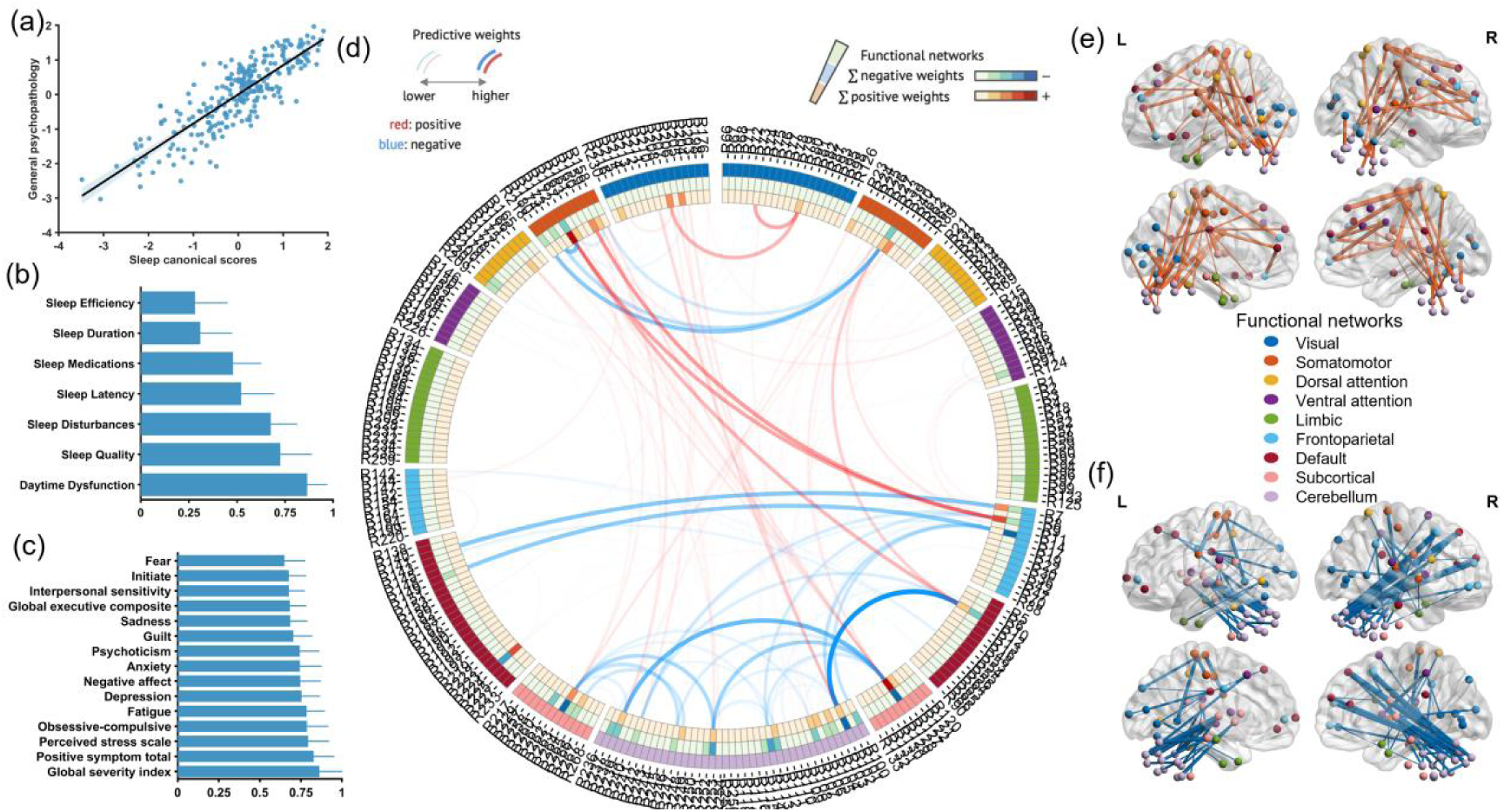
First canonical dimension linking broad sleep disturbance with general symptom burden and its connectome-wide expression. (a) Association between sleep and symptom canonical scores for the first dimension. Each point represents one participant, and the fitted line shows the linear association between the two canonical variates. (b) Canonical structure coefficients for the Pittsburgh Sleep Quality Index components. Positive coefficients indicate that higher expression of this dimension was associated with greater subjective sleep disturbance. The first dimension was most strongly characterized by daytime dysfunction, together with poorer subjective sleep quality, greater sleep disturbance, longer sleep latency and greater use of sleep medication. (c) Canonical structure coefficients for symptom-related variables. Higher expression of this dimension was associated with broader transdiagnostic symptom burden, including higher global severity, perceived stress, fatigue, depression, anxiety, obsessive-compulsive symptoms, negative affect and psychoticism. Asterisks indicate loadings meeting the prespecified stability or significance criterion. (d) Circular connectome representation of functional connectivity associations with the first canonical dimension. Edges indicate connections associated with individual expression of the dimension, with positive and negative associations shown separately according to the colour scale. Outer rings summarize node-level contributions across functional networks. (e) Positive subnetwork identified by network-based statistics. Higher expression of the first dimension was associated with stronger cross-network connectivity mainly involving somatomotor and higher-order systems, including somatomotor-frontoparietal and somatomotor-default mode connections. (f) Negative subnetwork identified by network-based statistics. Higher expression of the first dimension was associated with lower connectivity linking the default mode network with frontoparietal and cerebellar systems, together with lower cerebellar-subcortical connectivity. Network labels follow the visual, somatomotor, dorsal attention, ventral attention, limbic, frontoparietal and default mode networks, with additional subcortical and cerebellar systems comprising subcortical and cerebellar parcels, respectively.

**Fig. 2.**
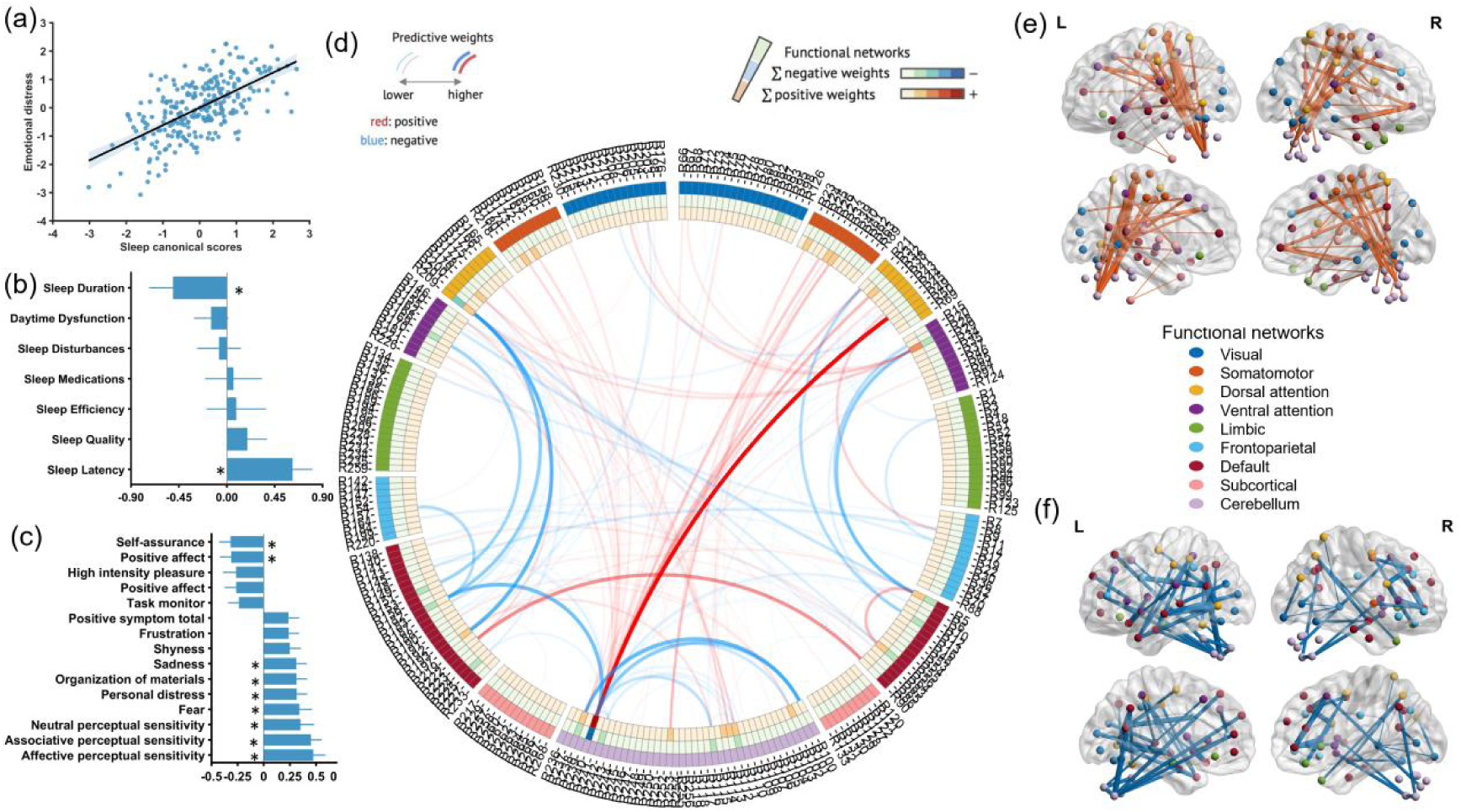
**Second canonical dimension linking shorter sleep duration and prolonged sleep latency with affective and perceptual sensitivity**. (a) Association between sleep and symptom canonical scores for the second dimension. Each point represents one participant, and the fitted line shows the linear association between the two canonical variates. (b) Canonical structure coefficients for the Pittsburgh Sleep Quality Index components. Higher expression of this dimension was primarily associated with shorter sleep duration and longer sleep latency, with weaker contributions from other sleep components. (c) Canonical structure coefficients for symptom-related variables. This dimension was characterized by lower positive affective and self-related disposition, including lower self-assurance and positive affect, together with greater affective, interpersonal and perceptual sensitivity. Asterisks indicate loadings meeting the prespecified stability or significance criterion. (d) Circular connectome representation of functional connectivity associations with the second canonical dimension. Edges indicate connections associated with individual expression of the dimension, with positive and negative associations shown separately according to the colour scale. Outer rings summarize node-level contributions across functional networks. (e) Positive subnetwork identified by network-based statistics. Higher expression of the second dimension was associated with stronger coupling between cerebellar and dorsal attention systems. (f) Negative subnetwork identified by network-based statistics. Higher expression of the second dimension was associated with weaker coupling between the default mode network and attention or cerebellar systems. Network labels follow the visual, somatomotor, dorsal attention, ventral attention, limbic, frontoparietal and default mode networks, with additional subcortical and cerebellar systems comprising subcortical and cerebellar parcels, respectively.

### Dimension 1: broad sleep disturbance linked to general symptom burden

The first canonical dimension was characterised by broadly positive loadings across nearly all Pittsburgh Sleep Quality Index components. The strongest loading was observed for daytime dysfunction, followed by subjective sleep quality, sleep disturbances, sleep latency, and use of sleep medication (Fig. 1b). Sleep duration and sleep efficiency also contributed in the same direction, although with relatively smaller loadings. Taken together, the sleep side of this dimension was most consistent with broad subjective sleep disturbance, with particular emphasis on daytime dysfunction.

On the symptom side, the same dimension showed positive loadings across a wide range of transdiagnostic symptom-related measures. The largest contributors included Brief Symptom Inventory Global Severity Index, Brief Symptom Inventory Positive Symptom Total (reflecting the number of endorsed symptoms), and Perceived Stress Scale score, obsessive-compulsive symptoms, fatigue, depression, negative affect, anxiety, and psychoticism (Fig. 1c), together with additional contributions from executive and interpersonal difficulties. This pattern suggests that Dimension 1 captured shared variation between broad sleep disturbance and a general burden of transdiagnostic symptom-related features.

Accordingly, higher expression of this dimension reflected individuals who reported poorer sleep across multiple sleep domains, especially greater daytime dysfunction, together with higher overall levels of transdiagnostic symptom burden.

### Dimension 2: short sleep and prolonged sleep latency linked to affective sensitivity and reduced positive disposition

The second canonical dimension showed a more selective sleep profile. On the sleep side, the most prominent feature was a strong negative loading for sleep duration, indicating that higher scores on this dimension were associated with shorter sleep. Sleep latency loaded positively, indicating greater difficulty initiating sleep. The remaining Pittsburgh Sleep Quality Index components contributed comparatively weakly (Fig. 2b). Thus, unlike Dimension 1, this dimension did not reflect global sleep disturbance, but rather a more specific pattern centered on short sleep and prolonged sleep onset.

On the symptom side, Dimension 2 was characterized by a contrast between lower positive disposition and greater affective or perceptual sensitivity. Negative loadings were observed for self-assurance and positive affect. Positive loadings were observed for affective, associative, and neutral perceptual sensitivity, as well as for fear, personal distress, and sadness (Fig. 2c). This pattern suggests that Dimension 2 captured a dimension marked by shorter sleep and delayed sleep initiation in combination with lower positive affective tone and heightened emotional or perceptual sensitivity.

Higher expression of this dimension therefore reflected a more specific sleep phenotype involving reduced sleep duration and greater difficulty falling asleep, accompanied by greater sensitivity to affective and perceptual experiences and lower positive self-related or reward-related disposition.

### Dimension-specific connectome-wide signatures

#### Dimension 1 was associated with reduced default mode coupling and increased somatomotor-higher-order coupling

Network-based statistics identified both a significant positive subnetwork (116 edges) and a significant negative subnetwork (132 edges) associated with the canonical score for Dimension 1 (pFWE<0.05) (Fig. 1d).

The negative subnetwork was concentrated in connections linking the default mode network with frontoparietal and cerebellar systems, together with reduced connectivity between cerebellar and subcortical networks (Fig. 1f). In other words, higher expression of the broad sleep disturbance and general symptom burden dimension was associated with lower connectivity among internally oriented, executive-control, cerebellar, and subcortical regulatory systems.

By contrast, the positive subnetwork showed increased cross-network connectivity involving somatomotor and higher-order systems (Fig. 1e), including somatomotor-frontoparietal and somatomotor-default mode connections. At a descriptive level, Dimension 1 was therefore associated with weaker default mode coupling with frontoparietal and cerebellar systems, alongside stronger coupling between somatomotor and higher-order networks (Table S5).

#### Dimension 2 was associated with a distinct pattern involving attention, default mode, and cerebellar systems

Network-based statistics identified both a significant positive subnetwork (124 edges) and a significant negative subnetwork (103 edges) associated with Dimension 2 (pFWE < 0.05; Fig. 2d), but the topography differed from that observed for Dimension 1.

The positive subnetwork for Dimension 2 primarily involved stronger coupling between cerebellar and dorsal attention systems (Fig. 2e). This pattern suggests that greater expression of the short sleep and prolonged sleep latency dimension was associated with stronger cerebellar-attention connectivity. The negative subnetwork for Dimension 2 was more prominent in connections linking default mode regions with attention and cerebellar systems (Fig. 2f). Thus, Dimension 2 was associated with weaker coupling between the default mode network and attention or cerebellar systems, alongside stronger coupling between cerebellar and dorsal attention systems (Table S5).

Taken together, the connectome-wide findings indicate that the two latent sleep-symptom dimensions were expressed in distinct patterns of large-scale functional connectivity rather than a common network configuration.

### Control analyses results

We conducted a series of control analyses to assess the robustness and generalizability of the canonical dimensions. LC1 showed consistent loadings across the control analyses, with correlations between the control and original solutions ranging from 0.73 to 0.99 for sleep loadings and from 0.94 to 0.99 for symptom loadings (Table S4). In the 5-fold cross-validation analysis, LC1 also showed significant correspondence between the held-out and original solutions (mean r = 0.65, *p* = 0.001), supporting its robustness and generalizability.

LC2 showed weaker and more variable correspondence. Correlations with the original solution ranged from 0.72 to 0.99 for sleep loadings and from 0.52 to 0.99 for symptom loadings (Table S4). The lower correlations were observed particularly in male participants (sleep r = 0.72; symptom r = 0.52) and healthy controls (sleep r = 0.74; symptom r = 0.60), although the corresponding correlations remained positive. Consistent with this reduced stability, LC2 did not show significant correspondence in the 5-fold cross-validation analysis (mean r = 0.20, p = 0.19). Together, these findings support the robustness of LC1 across analytic specifications. By contrast, the weaker cross-validation and subgroup correspondence for LC2 indicate that this dimension was less stable and should therefore be interpreted as a more circumscribed sleep-symptom profile requiring further validation.

## Discussion

The present study identified two latent dimensions linking multidimensional sleep characteristics with transdiagnostic symptom-related features and showed that these dimensions were associated with distinct connectome-wide patterns of resting-state functional connectivity. The first and more robust dimension reflected broad subjective sleep disturbance, particularly daytime dysfunction, together with general symptom burden. The second dimension reflected a more specific profile of shorter sleep duration and prolonged sleep latency, accompanied by lower positive disposition and greater affective and perceptual sensitivity. These findings support the view that sleep disturbance is transdiagnostic but not unitary [3,6,11,12]. They also extend dimensional models of psychopathology by showing that sleep-related variation can be organized into separable symptom-linked dimensions with distinct neural correlates [9,20,36,38,39].

The first dimension is best interpreted as a broad sleep-distress dimension. This pattern suggests that broad sleep disturbance may index a general burden state across diagnostic boundaries rather than a sleep signature of any specific disorder. It is consistent with evidence that sleep disturbances are common across mental disorders and with hierarchical accounts in which shared symptom variance reflects common liabilities cutting across diagnoses [6,9,12]. This result has direct implications for psychiatric research. Sleep disturbance is often treated as a secondary symptom of psychiatric disorders, a global severity marker, or a nuisance factor in neuroimaging studies. Our findings suggest a more specific view that sleep may constitute a functional dimension of general psychopathology. In particular, daytime dysfunction may be a key bridge between sleep complaints and psychiatric symptoms, because it captures the extent to which poor sleep is experienced as impaired alertness, reduced energy, and difficulty meeting daily demands. This interpretation is consistent with growing evidence that the daytime consequences of poor sleep are closely linked to psychiatric symptom burden and impaired daily functioning [40], and with evidence that different sleep domains have distinct clinical relevance [12,13,15,16]. It also helps explain why studies relying on global sleep indices may yield inconsistent findings: total scores can collapse separable sleep-symptom processes into a single summary measure [13,27,28]. The connectivity pattern of the first dimension provides a plausible neural context for this broad burden profile. Higher expression of this dimension was associated with reduced coupling among default mode, frontoparietal, cerebellar, and subcortical systems, together with increased coupling between somatomotor and higher-order networks. Dimension 1 was characterized by a broader disruption of large-scale network integration involving systems supporting internal cognition, executive regulation, cerebellar coordination, and affective-arousal control. This pattern suggests that the neural architecture underlying broad sleep-related functional impairment and transdiagnostic symptom burden may involve dysregulated interactions across multiple regulatory systems rather than dysfunction within isolated networks. Reduced DMN-FPN coupling may reflect impaired coordination between internally oriented mentation and executive control processes that support flexible transitions between self-generated thoughts and goal-directed behavior. The DMN and FPN dynamically interact to balance internally oriented cognition with externally directed cognitive control, supporting flexible adaptation to environmental demands [41,42]. Therefore, weakened integration between these systems may indicate reduced efficiency in aligning internal cognitive states with goal-directed control processes. Given that daytime dysfunction showed the strongest contribution on the sleep side of this dimension, disrupted DMN-FPN integration may represent one potential neural mechanism underlying difficulties in maintaining cognitive regulation and adaptive functioning in daily life. Reduced DMN-cerebellar coupling may additionally indicate diminished cerebellar support for organizing, monitoring, and refining internally generated cognition. The cerebellum is increasingly recognized as a component of distributed cognitive and affective regulatory circuits, contributing to predictive processing, internal model formation, and adaptive regulation beyond the motor domain [43–46]. Weakened cerebellar-subcortical connectivity may further reflect reduced coordination between cerebellar regulatory processes and broader subcortical systems involved in cognition and adaptive regulation [47,48]. Together, these findings suggest weakened coordination among cortical, cerebellar, and subcortical systems supporting cognitive regulation, predictive processing, and adaptive behavioral control, providing a potential neural substrate linking sleep-related functional impairment with transdiagnostic psychopathological burden.

In contrast, increased somatomotor coupling with higher-order networks suggests altered integration of bodily or sensorimotor representations with systems supporting cognition and self-related processing [41,49]. This enhanced coupling may reflect a compensatory-like reorganization in response to disrupted integration within core regulatory networks [50]. The concurrent weakening of DMN-FPN and DMN-cerebellar connectivity and strengthening of SMN-DMN and SMN-FPN coupling raises the possibility that somatomotor systems may be recruited to support integration when connectivity within core higher-order networks is reduced. Such reorganization could represent an adaptive attempt to maintain cognitive and behavioral functioning under conditions of widespread network disruption. However, this attempt may be limited in its ability to restore the integrity of core regulatory networks and may potentially increase processing demands, thereby contributing to the daytime functional difficulties. Importantly, because this dimension emerged from symptom measures spanning multiple clinical and affective domains rather than from a single diagnostic category, its distributed connectivity pattern may represent a neural correlate of shared psychopathological burden rather than disorder-specific dysfunction.

The second dimension showed a more selective profile. It was defined primarily by shorter sleep duration and longer sleep latency, with weaker contributions from other sleep components. Its symptom profile was characterized by lower self-assurance and positive affect, together with greater affective, interpersonal and perceptual sensitivity. The link between short sleep and psychological distress is consistent with prior epidemiological work [25], whereas the association with reduced positive affect is consistent with evidence that positive emotional dysfunction cuts across mood, anxiety and psychosis-spectrum conditions [51], and that sleep loss can attenuate positive affective functioning [52,53].

Unlike Dimension 1, which reflected broader disruption of network integration, Dimension 2 exhibited a more selective DMN-centered reorganization. Increased within-DMN connectivity accompanied by reduced DMN coupling with attentional and cerebellar systems suggests greater coherence of internally oriented processing together with weakened coordination with systems supporting external monitoring and predictive regulation [41]. Reduced DMN coupling with both DAN and VAN may indicate diminished integration between self-generated cognitive representations and attention-related systems involved in goal-directed allocation and stimulus-driven reorienting [54], potentially reducing flexibility in transitioning between internally focused cognition and externally directed monitoring. This imbalance may be particularly relevant to prolonged sleep latency, as difficulty disengaging from internally generated cognition and reducing pre-sleep cognitive arousal are central to impaired sleep initiation [55]. Reduced coupling between the default mode network and cerebellar systems may further indicate disrupted coordination between internally oriented processing and cerebellar systems supporting predictive processing and adaptive regulation, potentially limiting the transition from sustained cognitive engagement to the lower-arousal state required for sleep initiation [48].

In contrast, stronger coupling between cerebellar and dorsal attention systems suggests a selective redistribution of cerebellar connectivity toward attentional systems rather than generalized cerebellar alteration. This pattern may reflect enhanced coupling between cerebellar predictive processes and goal-directed attentional systems, potentially supporting persistent attentional-predictive engagement that becomes maladaptive when sustained during periods requiring disengagement for sleep initiation. Consistent with this interpretation, recent behavioral evidence suggests that individuals with insomnia have difficulty disengaging attention from sleep-related stimuli, with greater disengagement difficulty associated with greater insomnia severity through heightened arousal [56,57]. Although narrower than Dimension 1, this profile may also have transdiagnostic relevance because its symptom pattern cuts across affective, interpersonal, and perceptual domains.

Collectively, Dimension 2 may reflect a sleep-initiation and sensitivity profile in which shorter sleep and prolonged sleep latency are accompanied by altered coordination among default mode, attention, and cerebellar systems. This configuration may be relevant to persistent attentional-predictive engagement and difficulty disengaging during sleep initiation, although this interpretation remains tentative because the present study did not directly measure physiological arousal, sleep-onset physiology, or attentional bias. Our findings extend cognitive models of insomnia by suggesting that persistent attentional engagement may reflect altered coordination between large-scale networks supporting attention, prediction, and internally oriented regulation.

The difference in robustness between the two dimensions is important. The first dimension showed reliable cross-validated generalization, supporting its interpretation as a stable broad sleep-distress dimension. In contrast, the second dimension did not generalize reliably across folds. This reduced generalizability does not preclude the potential relevance of LC2, but it indicates that this dimension should be interpreted cautiously and tested in larger independent samples. LC2 may reflect a narrower sleep phenotype, lower reliability of subjective sleep duration and sleep latency measures, or reduced power to estimate a smaller latent component. Future studies combining subjective sleep reports with actigraphy, polysomnography and longitudinal follow-up will be needed to determine whether this short sleep and sleep-latency profile predicts insomnia persistence, affective symptoms, or treatment response.

Together, the two dimensions clarify the role of sleep in transdiagnostic psychiatry. Sleep is not merely a secondary symptom shared by multiple disorders; it may organize clinically meaningful variation across symptoms, daytime functioning and large-scale brain networks. At the same time, sleep should not be treated as a single global construct. Broad sleep disturbance and daytime dysfunction appear to relate to general psychopathological burden, whereas short sleep and prolonged latency may relate to sensitivity-related vulnerability. A multidimensional sleep assessment can therefore reveal sleep-symptom pathways that would be obscured by total Pittsburgh Sleep Quality Index scores or single sleep parameters. This view also supports the clinical relevance of sleep as a modifiable transdiagnostic target [58], consistent with evidence that cognitive behavioral therapy for insomnia can improve sleep and reduce depressive and anxiety symptoms [59].

These findings can also be situated within broader multidimensional models of sleep health, such as the Ru-SATED framework [18]. Several PSQI components used here overlap conceptually with Ru-SATED domains, including satisfaction, efficiency, duration and daytime alertness. From this perspective, LC1, which was dominated by daytime dysfunction and broad subjective sleep complaints, may capture the waking consequences and perceived burden of poor sleep, whereas LC2 was more closely aligned with sleep duration and sleep-initiation difficulties. This correspondence reinforces the view that transdiagnostic sleep-psychopathology associations are better understood through separable sleep-health dimensions than through a single global index of poor sleep.

Several limitations should be noted. First, the cross-sectional design prevents causal inference. Longitudinal studies are needed to determine whether these sleep-symptom dimensions predict later symptom worsening or whether psychiatric symptoms drive subsequent sleep disturbance. Second, sleep was assessed using self-report. Although subjective sleep complaints are clinically important, future work should test whether similar dimensions emerge using actigraphy, polysomnography or ecological sleep measures, especially given the known discrepancy between subjective and objective sleep measures [60,61]. Third, although we decomposed the PSQI into its seven component scores, the PSQI does not capture the full range of sleep-health dimensions emphasized in broader frameworks such as Ru-SATED. In particular, it provides limited information about sleep regularity and circadian timing, and therefore cannot determine whether the day-to-day stability and timing of sleep-wake schedules contribute additional sleep-psychopathology dimensions. Future studies using actigraphy, sleep diaries or ecological sleep assessments will be needed to test whether regularity and timing show distinct associations with transdiagnostic symptom profiles and connectome-wide organization. Fourth, LC2 requires independent validation because it showed weaker subgroup correspondence and did not generalize reliably in cross-validation. Larger transdiagnostic samples with subjective and objective sleep measures will be needed to determine whether this short sleep and prolonged latency profile represents a stable sleep-symptom dimension. Fifth, the sample was transdiagnostic and heterogeneous in diagnosis and medication exposure. This heterogeneity is appropriate for identifying dimensions across diagnostic boundaries, but future studies should examine whether these dimensions vary by diagnosis, illness stage, medication status or treatment response.

## Conclusion

In conclusion, the present findings indicate that sleep-psychopathology associations are multidimensional. A robust broad sleep-distress dimension links daytime dysfunction and general symptom burden with altered connectivity among default mode, frontoparietal, cerebellar and somatomotor systems. A second, more circumscribed dimension links short sleep and prolonged sleep latency with lower positive disposition, heightened sensitivity and a distinct cerebellar-attention connectivity pattern. These results suggest that sleep may serve as a key transdiagnostic domain through which symptom burden, daily functioning and large-scale brain network organization are jointly expressed, while also motivating future work on sleep-health dimensions not directly captured by the PSQI, such as regularity and circadian timing.

## Materials and Methods

### Participants

Data were drawn from the YaleNeuroConnect dataset [37], a deeply phenotyped transdiagnostic fMRI cohort designed to support brain-behavior modeling across diagnostic boundaries and dimensional symptom domains. The original YaleNeuroConnect connectome release included 302 participants with diverse mental health profiles, including healthy controls and psychiatric participants. The majority of psychiatric participants (64.23%) had two or more comorbid diagnoses, with substantial overlap across diagnostic categories. This substantial diagnostic overlap makes the cohort particularly well suited for transdiagnostic investigation. Because diagnostic categories in the source dataset were non-exclusive and were reported for the full connectome sample, they were used here only to characterize the source cohort rather than to define diagnosis-specific analytic groups.

The final analytic sample included 260 participants, comprising 123 healthy controls and 137 psychiatric participants. Participants were included if they had complete resting-state functional magnetic resonance imaging data with visually confirmed whole-brain coverage and no artifacts or other quality issues, a maximum mean frame-to-frame displacement < 0.15 mm, fewer than 20% of volumes exceeding a framewise displacement of 0.2 mm [62], and complete sleep and symptom-related measures entered into the multivariate analyses. The dataset was collected at the Yale School of Medicine between 2018 and 2024. Participants were recruited through community advertisements and referrals from Yale-affiliated clinics, including the Yale Depression Program,

PRIME Research Clinic, and STEP Clinic. Written informed consent was obtained in accordance with protocols approved by the Yale Institutional Review Board (HIC #2000020891).

In addition to extensive neuroimaging data, participants underwent comprehensive behavioral and clinical characterization. The present study focused on resting-state fMRI data acquired during the resting-state runs of the YaleNeuroConnect protocol. Outside the scanner, participants underwent a comprehensive assessment battery covering standardized neuropsychological testing, self-reported symptoms, demographic characteristics, psychiatric history, and cognitive functioning. Demographic and clinical characteristics of the final analytic sample are shown in Table 1.

**Table 1.** Demographic and clinical characteristics of the final analytic sample.

| Variable | Total sample<br>(N=260) | Healthy controls<br>(N=123) | Psychiatric participants<br>(N=137) |
| --- | --- | --- | --- |
| Sample size, n | 260 | 123 | 137 |
| Age, years, mean (SD) | 30.70 (10.92) | 30.80 (11.86) | 30.61 (10.05) |
| Sex, female/male | 148/112 | 67/56 | 81/56 |
| Education, years, mean (SD) | 15.88 (3.07) | 16.50 (2.81) | 15.32 (3.20) |
| Any medication use, n (%) | 126 (48.5%) | 37 (30.1%) | 89 (65.0%) |
| **Diagnostic status, n** |  |  |  |
| No mental-health diagnosis | 123 | 123 | 0 |
| Major depressive episode (MDE) | 76 | 0 | 76 |
| Generalized anxiety disorder (GAD) | 60 | 0 | 60 |
| Manic episode | 1 | 0 | 1 |
| Bipolar disorder | 23 | 0 | 23 |
| Alcohol use disorder (AUD) | 23 | 0 | 23 |
| Substance use disorder (SUD) | 22 | 0 | 22 |
| Panic episode | 27 | 0 | 27 |
| Post-traumatic stress disorder (PTSD) | 21 | 0 | 21 |
| Social anxiety disorder (SAD) | 7 | 0 | 7 |
| Obsessive compulsive disorder (OCD) | 15 | 0 | 15 |
| Autism spectrum disorder (ASD) | 1 | 0 | 1 |
| Attention-deficit/ hyperactivity disorder (ADHD) | 30 | 0 | 30 |
| Agoraphobia | 10 | 0 | 10 |
| Psychotic episode/ schizophrenia | 19 | 0 | 19 |
| Borderline personality disorder (BPD) | 8 | 0 | 8 |
| **Race, n (%)** |  |  |  |
| White (not Hispanic origin) | 120 (46.2%) | 50 (40.7%) | 70 (51.1%) |
| Black (not Hispanic origin) | 42 (16.2%) | 24 (19.5%) | 18 (13.1%) |
| Biracial (not Hispanic origin) | 6 (2.3%) | 3 (2.4%) | 3 (2.2%) |
| Black (Hispanic) | 8 (3.1%) | 2 (1.6%) | 6 (4.4%) |
| White (Hispanic) | 27 (10.4%) | 11 (8.9%) | 16 (11.7%) |
| Biracial (Hispanic) | 6 (2.3%) | 2 (1.6%) | 4 (2.9%) |
| Asian or Pacific Islander | 40 (15.4%) | 27 (22.0%) | 13 (9.5%) |
| Native | 1 (0.4%) | 1 (0.8%) | 0 (0.0%) |
| Other | 8 (3.1%) | 2 (1.6%) | 6 (4.4%) |
| Multiple races selected | 2 (0.8%) | 1 (0.8%) | 1 (0.7%) |
| **Annual household income, n (%)** |  |  |  |
| \$0-\$19,999 | 57 (21.9%) | 22 (17.9%) | 35 (25.5%) |
| \$20,000-\$34,999 | 49 (18.8%) | 20 (16.3%) | 29 (21.2%) |
| \$35,000-\$49,999 | 44 (16.9%) | 21 (17.1%) | 23 (16.8%) |
| \$50,000-\$99,999 | 65 (25.0%) | 33 (26.8%) | 32 (23.4%) |
| \$100,000-\$249,999 | 38 (14.6%) | 23 (18.7%) | 15 (10.9%) |
| Over \$250,000 | 7 (2.7%) | 4 (3.3%) | 3 (2.2%) |
| **Employment status, n (%)** |  |  |  |
| Full time | 101 (38.8%) | 45 (36.6%) | 56 (40.9%) |
| Part time (regular hours) | 33 (12.7%) | 10 (8.1%) | 23 (16.8%) |
| Part time (irregular hours) | 24 (9.2%) | 11 (8.9%) | 13 (9.5%) |
| Student | 82 (31.5%) | 52 (42.3%) | 30 (21.9%) |
| Retired/disability | 3 (1.2%) | 1 (0.8%) | 2 (1.5%) |
| Unemployed | 10 (3.8%) | 2 (1.6%) | 8 (5.8%) |
| Residential setting | 1 (0.4%) | 0 (0.0%) | 1 (0.7%) |
| Multiple employment statuses | 6 (2.3%) | 2 (1.6%) | 4 (2.9%) |

**Note.** Values are mean (SD) or n (%). Healthy controls and psychiatric participants were defined according to the dataset-level mental-health diagnosis grouping variable in the final analytic dataset. Diagnosis-specific counts are reported for the final analytic sample and are non-exclusive because psychiatric participants could meet criteria for more than one diagnosis; therefore, counts may sum to more than the number of psychiatric participants. Race and employment status are reported as mutually exclusive categories; participants selecting more than one category were coded as multiple races selected or multiple employment statuses. Medication use indicates any reported medication use.

### Sleep measures

Sleep was characterized using the seven component scores of the Pittsburgh Sleep Quality Index [63] rather than the global total score. These components indexed subjective sleep quality, sleep latency, sleep duration, habitual sleep efficiency, sleep disturbances, use of sleep medication, and daytime dysfunction. Descriptive statistics for these sleep measures across the total sample and subgroups are provided in Table S3. Treating these dimensions separately allowed us to preserve heterogeneity across sleep domains and to test whether distinct aspects of sleep covaried with different symptom profiles.

### Symptom-related measures

We examined 55 transdiagnostic symptom-related variables derived from multiple clinical instruments (Table S1), including the Adult Temperament Questionnaire (ATQ), which assesses temperamental traits and personality [64]; the Behavior Rating Inventory of Executive Function-Adult (BRIEF-A), measuring a set of inter-related higher-order cognitive abilities involved in self-regulatory functions [65]; the Brief Symptom Inventory (BSI), used to reflect the typical symptomatology of people with psychological problems [66]; the Interpersonal Reactivity Index (IRI), which evaluates empathy as a multidimensional individual difference construct [67]; the Positive and Negative Affect Schedule-Expanded (PANAS-X), which employs a hierarchical taxonomic scheme to assess both broad dimensions and specific, distinguishable emotional states [68] and the Perceived Stress Scale (PSS), which measures the degree to which situations in one’s life are appraised as stressful [69]. Because these measures span clinical symptoms, affective traits, stress, executive difficulties, and interpersonal sensitivity, they were analyzed jointly as a broad transdiagnostic symptom-related feature space. Descriptive statistics for the symptom-related measures across the total sample and subgroups are provided in Table S2. Diagnostic categories were not entered into the canonical correlation analysis; they were used only to describe the transdiagnostic source cohort and to distinguish healthy controls from psychiatric participants in descriptive and sensitivity analyses.

### Imaging acquisition protocol

Imaging data were obtained from the Yale NeuroConnect dataset. MRI scans were acquired on a 3T Siemens Prisma scanner equipped with a 64-channel head coil. High-resolution structural images were acquired using a T1-weighted magnetization-prepared rapid acquisition gradient-echo (MPRAGE) sequence (TR = 2,400 ms, TE = 1.22 ms, flip angle = 8°, voxel size = 1 mm³). Functional images were collected using a multiband echo-planar imaging (EPI) sequence (TR = 1,000 ms, TE = 30 ms, flip angle = 55°, slice thickness = 2 mm, multiband factor = 5). Resting-state fMRI data were acquired during two runs, with participants instructed to rest with their eyes open while fixating on a central cross. Detailed descriptions of the imaging protocol have been reported previously in the Yale NeuroConnect dataset publication [37].

#### Functional MRI Data Processing and Resting-state functional connectivity construction

Functional MRI data were preprocessed using the YaleNeuroConnect processing pipeline described in the original dataset publication. Structural images underwent skull stripping using optiBET, and motion correction was performed using SPM12. Structural images were nonlinearly registered to MNI space, and functional images were linearly aligned to structural images using FSL (v6.0.1) and BioImage Suite. Denoising procedures included nuisance regression of mean white matter, cerebrospinal fluid, and gray matter signals, regression of 24 motion parameters, removal of linear, quadratic, and cubic drift terms, and low-pass Gaussian filtering (σ = 1.55). Subjects were included in the final analyses if they had a maximum mean frame-to-frame displacement below 0.2 mm and fewer than 20% of volumes exceeding a framewise displacement of 0.2 mm [62]. The brain was parcellated using the Shen 268-node atlas [70], and pairwise functional connectivity was computed between all parcels to generate a 268 × 268 functional connectivity matrix for each participant. Connectivity values were Fisher z-transformed prior to subsequent analyses.

For visualization and network-level description, parcels were assigned to Yeo’s seven canonical large-scale brain systems [71], including the visual network (VN), somatomotor network (SMN), dorsal attention network (DAN), ventral attention network (VAN), limbic network (LN), frontoparietal network (FPN), and default mode network (DMN). Subcortical and cerebellar parcels were treated as two additional systems and labelled as the subcortical network and cerebellar network, respectively.

### Canonical correlation analysis

To identify latent dimensions linking multidimensional sleep and transdiagnostic symptom-related measures, we performed canonical correlation analysis. Age, sex and years of education were regressed out from both the sleep and symptom-related variables prior to model fitting.

CCA identifies linear combinations of variables from two datasets that are maximally correlated. In the present study, one data matrix comprised the seven PSQI component scores across 260 participants (260 × 7), and the other comprised 55 symptom-related variables across the same participants (260 × 55). CCA then derives pairs of canonical variates—one for sleep and one for symptom-related features—such that each pair is maximally correlated. The number of canonical variates is determined by the number of sleep measures, yielding seven LCs in this analysis. Each LC is described by a pattern of weights for sleep measures and a corresponding pattern for symptom-related measures. Projecting the original data onto these weights produces participant-specific composite scores for both sleep and symptom-related features. The contribution of each original variable to an LC was quantified using Pearson correlations between the variable and its corresponding composite score, resulting in canonical structure coefficients. These structure coefficients reflect the association between each original variable and the corresponding canonical variate and were used to aid interpretation of each dimension. Statistical significance of the canonical modes was assessed using permutation testing (10,000 permutations) with FDR correction. Finally, the stability of the loadings was evaluated using bootstrap resampling (10,000 samples), which provided confidence intervals for each measure’s loading. Based on these tests, two canonical dimensions were retained for interpretation.

### Connectome-wide analysis of canonical dimensions

To determine whether the identified sleep-symptom dimensions mapped onto distinct patterns of resting-state connectivity, we performed connectome-wide analyses using network-based statistics. Specifically, generalized linear models were used to relate each participant’s dimension-specific canonical score, computed as the average of the sleep and symptom canonical scores [72], to resting-state functional connectivity, after controlling for age, sex, and education.

Network-based statistics [73] were then used to identify connected components showing significant positive or negative associations with the canonical score. The primary edge-forming threshold was set at p <0.001, and component significance was assessed using 10,000 permutations with family-wise error correction, considering components significant at pFWE < 0.05.

### Control analyses

We conducted several control analyses to assess the robustness of our findings. First, we performed 5-fold cross-validation to evaluate the generalizability of the sleep-symptom profiles. In each fold, a CCA model was trained on 80% of the participants and applied to the remaining 20%. Canonical coefficients from the training set were projected onto the test set to obtain sleep and symptom scores, and Pearson correlations between these scores were computed. Statistical significance of the cross-validated correlations was assessed using permutation testing (10,000 permutations). Second, we assessed the influence of covariates on the identified sleep-symptom profiles without removing their effects, and further evaluated the potential contributions of race and income by including them as covariates in the analyses. Third, to reduce potential differences in scale across variables, we repeated the CCA after applying quantile normalization to both the sleep and symptom measures. Fourth, we repeated the CCA separately within healthy controls and psychiatric participants, as defined by the dataset-level mental-health diagnosis grouping variable, to examine whether the identified sleep-symptom associations were consistent within each population. Fifth, we repeated the CCA separately within female and male participants to examine whether the sleep-symptom associations were consistent across sexes. Sixth, we conducted the CCA exclusively in participants who were not taking any medications to ensure that the identified sleep-symptom associations were not influenced by pharmacological effects. To assess robustness, we calculated the Pearson correlations between the sleep and symptom loadings derived from each control analysis and the corresponding loadings from the original CCA. All post hoc comparisons were corrected for multiple testing using the false discovery rate procedure, with a threshold of q < 0.05.

## Data Availability

Data from the Yale NeuroConnect dataset is publicly available at https://yaleedu-my.sharepoint.com/:f:/g/personal/todd_constable_yale_edu/En99p1wBK6xKj3Zuh2T48_cBVN_oUWEoDlDoVjKhZiMXtQ. Code for the CCA is available at https://github.com/valkebets/sleep_biopsychosocial_profiles. The Network-Based Statistic (NBS) software is available at https://www.nitrc.org/projects/nbs/.

https://github.com/valkebets/sleep_biopsychosocial_profiles

## Acknowledgements

We thank the participants and investigators involved in the Yale NeuroConnect dataset for making this resource available for secondary analyses.

## Data and code availability

Data from the Yale NeuroConnect dataset is publicly available at https://yaleedu-my.sharepoint.com/:f:/g/personal/todd_constable_yale_edu/En99p1wBK6xKj3Zuh 2T48_cBVN_oUWEoDlDoVjKhZiMXtQ. Code for the CCA is available at https://github.com/valkebets/sleep_biopsychosocial_profiles. The Network-Based Statistic (NBS) software is available at https://www.nitrc.org/projects/nbs/.

## Author Contributions

**Conceptualization:** Yishan Kou, Yulin Wang

**Data curation:** Xueer Jin, Debo Dong

**Formal analysis:** Debo Dong, Yulin Wang, Yishan Kou

**Funding Acquisition:** Yulin Wang

**Methodology:** Debo Dong, Yulin Wang

**Visualization:** Debo Dong

**Writing – Original Draft Preparation:** Yishan Kou

**Writing – Review & Editing:** Yishan Kou, Xueer Jin, Xu Lei, Yulin Wang, Debo Dong

