## Supplementary Materials (PDF) for "Latent transdiagnostic dimensions linking sleep and psychopathology map onto distinct connectome-wide networks"

### Supplementary Methods

**Table S1. Description of psychological measures included in the transdiagnostic symptom-related feature space** Note. Adult Temperament Questionnaire, ATQ [1]; Behavior Rating Inventory of Executive Function, BRIEF [2]; Brief Symptom Inventory, BSI [3]; Interpersonal Reactivity Index, IRI [4]; Positive and Negative Affect Schedule, PANAS [5]; the Perceived Stress Scale, PSS [6].

| Measure | Items and scoring | Dimensions/subscales |
|---|---|---|
| Adult Temperament Questionnaire (ATQ) | 77 items; rated on a 7-point Likert scale | Activation control, Effortful attention, Inhibitory control, Affective perceptual sensitivity, Neutral perceptual sensitivity, Associative perceptual sensitivity, Frustration, Fear, Sadness, Discomfort, High intensity pleasure, Positive affect, Sociability |
| Behavior Rating Inventory of Executive Function–Adult (BRIEF-A) | 75 items; 3-point response scale | Behavioral regulation index, Inhibit, Shift, Emotional control, Metacognition index, Initiate, Working memory, Plan/organize, Organization of materials, Task monitor, Self-monitor, Global executive composite |
| Brief Symptom Inventory (BSI) | 53 items; 5-point Likert scale | Somatization, Obsessive-compulsive, Interpersonal sensitivity, Depression, Anxiety, Hostility, Phobic anxiety, Paranoid ideation, Psychoticism, Global severity index, Positive symptom total, Positive symptom distress index |
| Interpersonal Reactivity Index (IRI) | 28 items; 5-point Likert scale | Fantasy, Empathic concern, Perspective taking, Personal distress |
| Positive and Negative Affect Schedule–Expanded (PANAS-X) | 60 items; 5-point Likert scale | Negative affect, Fear, Sadness, Guilt, Hostility, Shyness, Fatigue, Positive affect, Joviality, Self-assurance, Attentiveness, Serenity, Surprise |
| Perceived Stress Scale (PSS) | 10 items; 5-point Likert scale | Perceived stress scale |

#### Supplementary Results

**Table S2. Descriptive statistics for symptom-related measures across total sample and subgroups (M ± SD)** Note. M = mean; SD = standard deviation. N = sample size per group. Adult Temperament Questionnaire, ATQ [1]; Behavior Rating Inventory of Executive Function, BRIEF [2]; Brief Symptom Inventory, BSI [3]; Interpersonal Reactivity Index, IRI [4]; Positive and Negative Affect Schedule, PANAS [5]; the Perceived Stress Scale, PSS [6].

| Item number | Assessment | Measure | Total Sample (N=260) | Healthy Controls (N=123) | Patients (N=137) |
|---|---|---|---|---|---|
| 1 | ATQ | Activation control | 4.82 ± 1.05 | 5.17 ± 0.92 | 4.50 ± 1.07 |
| 2 | ATQ | Effortful attention | 4.16 ± 1.27 | 4.59 ± 1.23 | 3.77 ± 1.18 |
| 3 | ATQ | Inhibitory control | 4.54 ± 0.99 | 4.80 ± 0.93 | 4.31 ± 0.99 |
| 4 | ATQ | Affective perceptual sensitivity | 4.70 ± 1.17 | 4.49 ± 1.19 | 4.89 ± 1.12 |
| 5 | ATQ | Neutral perceptual sensitivity | 4.73 ± 1.14 | 4.71 ± 1.08 | 4.75 ± 1.20 |
| 6 | ATQ | Associative perceptual sensitivity | 4.59 ± 1.27 | 4.10 ± 1.20 | 5.02 ± 1.17 |
| 7 | ATQ | Frustration | 3.54 ± 1.08 | 3.31 ± 1.02 | 3.75 ± 1.09 |
| 8 | ATQ | Fear | 3.54 ± 1.06 | 3.13 ± 0.85 | 3.91 ± 1.09 |
| 9 | ATQ | Sadness | 4.28 ± 1.11 | 3.94 ± 0.94 | 4.57 ± 1.16 |
| 10 | ATQ | Discomfort | 3.86 ± 1.36 | 3.63 ± 1.32 | 4.08 ± 1.36 |
| 11 | ATQ | High intensity pleasure | 4.12 ± 1.13 | 4.06 ± 1.12 | 4.18 ± 1.13 |
| 12 | ATQ | Positive affect | 4.31 ± 1.25 | 4.82 ± 0.99 | 3.86 ± 1.28 |
| 13 | ATQ | Sociability | 4.45 ± 1.30 | 4.76 ± 1.18 | 4.17 ± 1.35 |
| 14 | BRIEF | Behavioral regulation index | 44.56 ± 11.44 | 39.50 ± 9.13 | 49.10 ± 11.43 |
| 15 | BRIEF | Inhibit | 12.15 ± 3.39 | 10.76 ± 2.85 | 13.40 ± 3.35 |
| 16 | BRIEF | Shift | 9.34 ± 2.74 | 8.24 ± 2.18 | 10.34 ± 2.82 |
| 17 | BRIEF | Emotional control | 14.77 ± 4.91 | 12.80 ± 3.58 | 16.53 ± 5.26 |
| 18 | BRIEF | Metacognition index | 62.26 ± 15.69 | 55.59 ± 13.13 | 68.24 ± 15.42 |
| 19 | BRIEF | Initiate | 12.84 ± 3.74 | 11.20 ± 2.94 | 14.31 ± 3.77 |
| 20 | BRIEF | Working memory | 12.75 ± 4.02 | 11.17 ± 3.33 | 14.16 ± 4.08 |
| 21 | BRIEF | Plan/organize | 14.95 ± 4.12 | 13.41 ± 3.42 | 16.34 ± 4.22 |
| 22 | BRIEF | Organization of materials | 12.20 ± 3.64 | 11.06 ± 3.10 | 13.23 ± 3.79 |
| 23 | BRIEF | Task monitor | 9.54 ± 2.46 | 8.76 ± 2.26 | 10.25 ± 2.42 |
| 24 | BRIEF | Self-monitor | 8.33 ± 2.54 | 7.66 ± 2.27 | 8.93 ± 2.62 |
| 25 | BRIEF | Global executive composite | 106.82 ± 25.32 | 95.10 ± 21.05 | 117.34 ± 24.25 |
| 26 | BSI | Somatization | 0.43 ± 0.95 | 0.28 ± 1.18 | 0.57 ± 0.65 |
| 27 | BSI | Obsessive-compulsive | 1.16 ± 1.01 | 0.67 ± 0.70 | 1.59 ± 1.06 |
| 28 | BSI | Interpersonal sensitivity | 0.85 ± 0.94 | 0.46 ± 0.63 | 1.20 ± 1.03 |
| 29 | BSI | Depression | 0.94 ± 1.13 | 0.33 ± 0.50 | 1.49 ± 1.26 |
| 30 | BSI | Anxiety | 0.67 ± 0.80 | 0.27 ± 0.41 | 1.04 ± 0.88 |
| 31 | BSI | Hostility | 0.48 ± 0.64 | 0.27 ± 0.41 | 0.66 ± 0.75 |
| 32 | BSI | Phobic anxiety | 0.37 ± 0.61 | 0.12 ± 0.27 | 0.60 ± 0.73 |
| 33 | BSI | Paranoid ideation | 0.55 ± 0.72 | 0.25 ± 0.36 | 0.81 ± 0.85 |
| 34 | BSI | Psychoticism | 0.56 ± 0.73 | 0.20 ± 0.34 | 0.89 ± 0.82 |
| 35 | BSI | Global severity index | 0.67 ± 0.65 | 0.31 ± 0.33 | 1.00 ± 0.70 |
| 36 | BSI | Positive symptom total | 17.75 ± 12.94 | 10.63 ± 9.38 | 24.15 ± 12.37 |
| 37 | BSI | Positive symptom distress index | 1.65 ± 2.20 | 1.43 ± 3.11 | 1.85 ± 0.66 |
| 38 | IRI | Fantasy | 16.16 ± 5.60 | 15.39 ± 5.80 | 16.85 ± 5.34 |
| 39 | IRI | Empathic concern | 20.95 ± 4.42 | 20.65 ± 4.52 | 21.21 ± 4.33 |
| 40 | IRI | Perspective taking | 19.52 ± 4.58 | 19.49 ± 4.58 | 19.54 ± 4.59 |

| Item number | Assessment | Measure | Total Sample (N=260) | Healthy Controls (N=123) | Patients (N=137) |
|---|---|---|---|---|---|
| 41 | IRI | Personal distress | 10.33 ± 5.07 | 8.98 ± 4.46 | 11.53 ± 5.28 |
| 42 | PANAS | Negative affect | 18.00 ± 7.45 | 14.79 ± 5.62 | 20.89 ± 7.71 |
| 43 | PANAS | Fear | 10.38 ± 4.95 | 8.47 ± 3.61 | 12.09 ± 5.37 |
| 44 | PANAS | Sadness | 9.96 ± 5.10 | 7.54 ± 3.41 | 12.13 ± 5.39 |
| 45 | PANAS | Guilt | 10.47 ± 5.79 | 8.08 ± 3.47 | 12.61 ± 6.57 |
| 46 | PANAS | Hostility | 9.47 ± 3.66 | 8.22 ± 2.95 | 10.59 ± 3.88 |
| 47 | PANAS | Shyness | 6.38 ± 2.71 | 5.65 ± 2.11 | 7.04 ± 3.01 |
| 48 | PANAS | Fatigue | 11.07 ± 3.98 | 9.67 ± 3.36 | 12.32 ± 4.09 |
| 49 | PANAS | Positive affect | 31.70 ± 8.30 | 34.77 ± 6.95 | 28.93 ± 8.45 |
| 50 | PANAS | Joviality | 24.18 ± 7.50 | 27.59 ± 5.82 | 21.12 ± 7.53 |
| 51 | PANAS | Self-assurance | 15.85 ± 5.44 | 17.45 ± 4.74 | 14.42 ± 5.65 |
| 52 | PANAS | Attentiveness | 13.31 ± 3.34 | 14.24 ± 2.95 | 12.47 ± 3.46 |
| 53 | PANAS | Serenity | 9.34 ± 3.08 | 10.31 ± 2.66 | 8.47 ± 3.19 |
| 54 | PANAS | Surprise | 5.72 ± 2.36 | 5.98 ± 2.54 | 5.49 ± 2.17 |
| 55 | PSS | Perceived stress scale | 22.46 ± 9.41 | 17.18 ± 7.31 | 27.20 ± 8.53 |

**Table S3. Descriptive statistics for sleep measures across total sample and subgroups (M ± SD)** Note. M = mean; SD = standard deviation. N = sample size per group. Pittsburgh Sleep Quality Index, PSQI [7].

| Item number | Assessment | Measure | Total Sample (N=260) | Healthy Controls (N=123) | Patients (N=137) |
|---|---|---|---|---|---|
| 1 | PSQI | Sleep quality | 1.08 ± 0.74 | 0.85 ± 0.63 | 1.30 ± 0.76 |
| 2 | PSQI | Sleep latency | 1.37 ± 0.97 | 0.95 ± 0.80 | 1.74 ± 0.96 |
| 3 | PSQI | Sleep duration | 0.73 ± 0.84 | 0.63 ± 0.75 | 0.83 ± 0.90 |
| 4 | PSQI | Habitual sleep efficiency | 0.50 ± 0.87 | 0.32 ± 0.70 | 0.66 ± 0.96 |
| 5 | PSQI | Sleep disturbances | 1.16 ± 0.58 | 0.99 ± 0.52 | 1.31 ± 0.60 |
| 6 | PSQI | Use of sleep medication | 0.61 ± 1.04 | 0.25 ± 0.66 | 0.93 ± 1.21 |
| 7 | PSQI | Daytime dysfunction | 0.94 ± 0.80 | 0.63 ± 0.68 | 1.23 ± 0.79 |

**Table S4. Correlations between sleep and symptom loadings from control analyses and those from the original analysis.**

| | | Not regressing out confounds | Further regressing out race and income | Quantile normalization | Female participants only | Male participants only | within the healthy control group | within the patient group | not taking any medications |
|---|---|---|---|---|---|---|---|---|---|
| LC1 | Sleep loadings | 0.99 | 0.99 | 0.99 | 0.96 | 0.95 | 0.73 | 0.96 | 0.92 |
| | Symptom loadings | 0.99 | 0.99 | 0.98 | 0.98 | 0.99 | 0.94 | 0.96 | 0.96 |
| LC2 | Sleep loadings | 0.99 | 0.99 | 0.99 | 0.95 | 0.72 | 0.74 | 0.87 | 0.85 |
| | Symptom loadings | 0.99 | 0.99 | 0.96 | 0.86 | 0.52 | 0.60 | 0.86 | 0.65 |

**Table S5. Summary of dominant network-pair connectivity patterns**

| Dimension | Subnetwork | Top network pairs by edge count |
|---|---|---|
| LC1 | positive | SMN-FPN, SMN-DMN |
| LC1 | negative | DMN-cerebellar, DMN-FPN, cerebellar-SCN |
| LC2 | positive | DAN-cerebellar |
| LC2 | negative | DMN-DAN, DMN-cerebellar, DMN-VAN |
